# In-depth virological assessment of kidney transplant recipients with COVID-19

**DOI:** 10.1101/2020.06.17.20132076

**Authors:** Ilies Benotmane, Gabriela Gautier Vargas, Marie-Josée Wendling, Peggy Perrin, Aurélie Velay, Xavier Bassand, Dimitri Bedo, Clement Baldacini, Mylene Sagnard, Dogan-Firat Bozman, Margaux Della Chiesa, Morgane Solis, Floriane Gallais, Noëlle Cognard, Jerome Olagne, Héloïse Delagreverie, Louise Gontard, Baptiste Panaget, David Marx, Francoise Heibel, Laura Braun, Bruno Moulin, Sophie Caillard, Samira Fafi-Kremer

## Abstract

Severe acute respiratory syndrome coronavirus 2 (SARS-CoV-2) has spread widely, causing coronavirus disease 2019 (COVID-19) and significant mortality. However, data on viral loads and antibody kinetics in immunocompromised populations are lacking. We aimed to determine nasopharyngeal and plasma viral loads via RT-PCR and SARS-CoV-2 serology via ELISA and study their association with severe forms of COVID-19 and death in kidney transplant recipients. In this study we examined hospitalized kidney transplant recipients with non-severe (n = 21) and severe (n =19) COVID-19. SARS-CoV-2 nasopharyngeal and plasma viral load and serological response were evaluated based on outcomes and disease severity. Ten recipients (25%) displayed persistent viral shedding 30 days after symptom onset. The SARS-CoV-2 viral load of the upper respiratory tract was not associated with severe COVID-19, whereas the plasma viral load was associated with COVID-19 severity (p=0.0087) and mortality (p=0.024). All patients harbored antibodies the second week after symptom onset that persisted for two months. We conclude that plasma viral load is associated with COVID-19 morbidity and mortality, whereas nasopharyngeal viral load is not. SARS-CoV-2 shedding is prolonged in kidney transplant recipients and the humoral response to SARS-CoV-2 does not show significant impairment in this series of transplant recipients.

## 1. Introduction

In December 2019, severe acute respiratory syndrome coronavirus 2 (SARS-CoV-2) emerged in China, and it has since spread widely across the world.^1^ The resulting coronavirus disease 2019 (COVID-19) has led to a high death toll. Scientific knowledge on SARS-CoV-2 has evolved rapidly since the outbreak, but little is known about responses to the virus in immunocompromised populations. Infection with respiratory viruses has been shown to be particularly concerning in transplant recipients due to prolonged viral shedding and a higher risk of complications.^2,3^ However, there have been no reports indicating whether SARS-CoV-2 infection presents the same risks for transplant recipients as other respiratory viruses. Determining viral loads and antibody kinetics in immunocompromised individuals is necessary to protect this highly vulnerable population. Because prolonged viral shedding and/or a lack of protective immunity could lead to significant viral spread in patients’ environments, protective measures may need to be increased.

We thus conducted a retrospective cohort study in kidney transplant recipients (KTR) in Alsace, Grand-Est France, to determine the dynamics of nasopharyngeal and plasma viral loads and SARS-CoV-2 serology and to study their association with mortality and severe forms of COVID-19.

## 2. Patients and Methods

### 2.1 Study population

A total of 40 adult KTR hospitalized with COVID-19 were recruited at our transplant center between March 4 and April 7, 2020. COVID-19 was diagnosed in these patients based on their clinical symptoms and positive reverse transcription-polymerase chain reaction (RT-PCR) results obtained using nasopharyngeal swabs and/or typical lung lesions observed through chest computed tomography (CT). Patient characteristics were retrieved from digital medical records from the day of admission through the date of last follow-up (May 13, 2020). Data for the following parameters were collected: demographic variables, symptoms and time of presentation, immunosuppressive therapy and management, laboratory parameters, chest CT findings, administered drugs, death, and intensive care unit (ICU) admissions. The study protocol was reviewed and approved by the local Institutional Review Board (approval number: DC-2013-1990).

### 2.2 Virological diagnosis and follow-up of SARS-CoV-2 infection

Quantitative RT-PCR tests for SARS-CoV-2 nucleic acid were performed using nasopharyngeal swabs and plasma samples obtained at admission and during follow-up for all but one patient, whose nasopharyngeal swab was collected at another laboratory at the time of diagnosis. Primer and probe sequences targeted two regions on the RNA-dependent RNA polymerase (*RdRp*) gene specific to SARS-CoV-2. The assay’s sensitivity was approximately 10 RNA copies per reaction. A SARS-CoV-2 RT-PCR result was considered positive when at least one of the targets was amplified.^4^

### 2.3 SARS-CoV-2 serological assessment

Immunoglobulin (Ig) M and IgG antibody responses to SARS-CoV-2 recombinant nucleocapsid and spike antigens were tested using a commercially available enzyme-linked immunosorbent assay (ELISA; DIA.PRO Diagnostic BioProbes Srl, Sesto San Giovanni, Italy) according to the manufacturer’s instructions. Antibody levels are presented as measured absorbance values divided by the cutoff (S/CO). The cutoff value was defined as the mean absorbance values of the three negative controls plus 0.250. IgM and IgG specificity and sensitivity reached an overall value of ≥ 98%. Seronegative patients were negative for IgM and IgG.

### 2.4 Statistical analysis

Continuous data are presented as medians and interquartile ranges (IQR) and were analyzed using the non-parametric Mann-Whitney *U* test. Categorical variables are expressed as counts and percentages and were compared using the Fisher’s exact test. The associations between maximum nasopharyngeal SARS-CoV-2 viral load and clinical, demographic, and laboratory variables were determined using Spearman’s correlation coefficient (ρ) values. Receiver operating characteristic (ROC) curves were generated to investigate the viral loads in the upper respiratory tract and plasma with respect to disease severity and mortality. Survival plots of severe COVID-19-free survival and COVID-19-specific survival were graphically represented with Kaplan-Meier curves according to RNAaemia (i.e., positive plasma viral load), using the log-rank test to compare differences in survival. Severe COVID-19 was defined as one or more of the following: an oxygen requirement of >6 L/min, the need for ICU admission, and patient death. Patients were censored at the time of last follow-up. Statistical analyses were performed using GraphPad Prism 8.0 (GraphPad Inc., San Diego, CA, USA). A p value <0.05 (two-tailed) was considered statistically significant.

## 3. Results

### 3.1 General characteristics of the study patients

A total of 40 patients were included in this study. Their demographic and clinical characteristics are presented in Table 1. The patients were mainly men (77.5%), with a median age of 63.8 years (IQR: 54.6−68.2 years) and a median body mass index (BMI) of 29.5 kg/m^2^ (IQR: 24−33 kg/m^2^). The median time after kidney transplantation was 6.6 years (IQR: 2.8−14.6 years). At the time of COVID-19 diagnosis, 35 (87.5%) patients were receiving calcineurin inhibitors (CNI). Mycophenolate mofetil (MMF) or mycophenolic acid (MPA) was being taken by 34 patients (85%), mTOR inhibitors were being administered to 6 (15%) patients, and steroids were taken by 23 (57.5%) patients. The median interval between the onset of symptoms and COVID-19 diagnosis was 4 days (IQR: 3−7 days). All but two patients had a fever. Respiratory (n=34, 85%) and gastrointestinal (n=31, 77.5%) symptoms were the most common clinical manifestations. Twenty-one patients had a non-severe clinical presentation, whereas 19 had a severe clinical course. During follow-up, the mortality rate was found to be 22.5% (9/40).

**Table 1.** Demographics and clinical characteristics of patients according to disease severity.

|  | **Hospitalized**<br>**patients (n=40)** | **Non-severe**<br>**patients (n=21)** | **Severe**<br>**patients**<br>**(n=19)** | **p** |
| --- | --- | --- | --- | --- |
| **Men** | 31 (77.5%) | 19 (90.5%) | 12 (63.1%) | **0.06** |
| **Age, years** | 63.8 [54.6-68.2] | 58.4 [50.9-64.3] | 65.5 [62.6-69.9] | **0.02** |
| **>60 years** | 25 (62.5%) | 9 (42.9%) | 16 (84.2%) | **0.01** |
| **Comorbidities** |  |  |  |  |
| **BMI, kg/m<sup>2</sup>** | 29.5 [24-33] | 25 [23-32] | 31 [27-33] | **0.07** |
| **<25** | 13 (32.5%) | 11 (52.4%) | 2 (10.5%) | **0.02** |
| **25-30** | 8 (20%) | 4 (19.1%) | 4 (21.1%) |  |
| **>30** | 20 (50%) | 7 (33.3%) | 13 (68.4%) |  |
| **Cardiovascular disease** | 16 (40%) | 8 (38.1%) | 8 (42.1%) | **1** |
| **Respiratory disease** | 9 (22.5%) | 5 (23.8%) | 4 (21%) | **1** |
| **Obstructive sleep apnea** | 7 (17.1%) | 4 (19.1%) | 3 (15%) | **1** |
| **Diabetes** | 19 (47.5%) | 8 (38.1%) | 11 (57.9%) | **0.34** |
| **Active cancer** | 0 | 0 | 0 |  |
| **Hypertension** | 33 (82.5%) | 15 (71.4%) | 18 (94.7%) | **0.09** |
| **RAAS inhibitor use** | 15 (37.5%) | 7 (33.3%) | 8 (42.1%) | **0.75** |
| **ACE inhibitor use** | 9 (22.5%) | 3 (14.3%) | 6 (31.6%) | **0.26** |
| **ARB use** | 6 (15%) | 4 (19.1%) | 2 (10.5%) | **0.66** |
| **Interval from kidney transplantation (years)** | 6.6 [2.8-14.6] | 3.8 [2.1-12.6] | 7.7 [5.2-14.9] | **0.22** |
| **Immunosuppressive therapy** |  |  |  |  |
| **Induction** |  |  |  |  |
| **immunosuppression** |  |  |  |  |
| **Anti-thymocyte globulin** | 18 (43.9%) | 10 (47.6%) | 8 (42.1%) | **0.9** |
| **Anti-CD25** | 19 (46.3%) | 9 (42.9%) | 10 (52.6%) |  |
| **No induction** | 3 (7.3%) | 2 (9.5%) | 1 (5%) |  |
| **Maintenance** |  |  |  |  |
| **immunosuppression** |  |  |  |  |
| **Tacrolimus** | 21 (52.5%) | 10 (47.6%) | 11 (57.9%) | **0.54** |
| **Ciclosporin** | 14 (35%) | 7 (33.3%) | 7 (36.8%) | **1** |
| **MMF/MPA** | 34 (85%) | 19 (90.5%) | 15 (78.9%) | **0.40** |
| **mTOR inhibitors** | 6 (15%) | 4 (19.1%) | 2 (10.5%) | **0.66** |
| **Azathioprine** | 1 (2.5%) | 0 | 1 (5.3%) | **0.47** |
| **Steroids** | 23 (57.5%) | 12 (57.1%) | 11 (57.9%) | **1** |
| **Belatacept** | 2 (5%) | 2 (9.5%) | 0 | **0.49** |
| **Eculizumab** | 1 (2.5%) | 0 | 1 (5.3%) | **0.47** |
| **Clinical symptoms during hospitalization** |  |  |  |  |
| **Dyspnea** | 28 (70%) | 9 (42.9%) | 19 (100%) | **<0.001** |
| **Cough** | 31 (77.5%) | 15 (71.4%) | 16 (84.2%) | **0.46** |
| **Fever** | 38 (95%) | 20 (95.2%) | 18 (94.7%) | **1** |
| **Myalgia** | 22 (55%) | 14 (66.7%) | 8 (42.1%) | **0.20** |
| **Headache** | 12 (30%) | 9 (42.8%) | 3 (15.8%) | **0.09** |
| **Diarrhea** | 31 (77.5%) | 19 (90.5%) | 12 (63.2%) | **0.06** |
| **Vomiting** | 7 (17.5%) | 5 (23.8%) | 2 (10.5%) | **0.41** |
| **Anosmia/ageusia** | 8 (20%) | 6 (28.6%) | 2 (10.5%) | **0.24** |
| **Neurological**<br>**manifestations** | 15 (37.5%) | 8 (38.1%) | 7 (36.9%) | **1** |

Continuous variables are presented as median (interquartile range), whereas categorical variables are presented as count (percentage).
Abbreviations: BMI, body mass index; RAAS, renin-angiotensin-aldosterone system; ACE, angiotensin converting enzyme; ARB, angiotensin receptor blockers; MMF, mycophenolate mofetil; MPA, mycophenolic acid, mTOR: mammalian target of rapamycin

### 3.2 Viral load in nasopharyngeal swabs

A total of 118 upper respiratory specimens were analyzed (median of three swabs per patient [IQR: 2−3 swabs]). The median viral load was 5.17 log^10^ copies/reaction (IQR: 3.80−6.69) at diagnosis (between day 0 and day 14 after symptom onset). A total of 29 patients (74.4%) had their peak viral load at admission, and two patients hospitalized at day 10 and day 11 after symptom onset had negative RT-PCR results at admission and during the follow-up. The viral load at admission was not statistically different between severe and non-severe patients (6.22 log^10^ copies/reaction *versus* 5.17 log^10^ copies/reaction, respectively p=0.29) and was not predictive of disease severity (area under the ROC curve = 0.595, p=0.31) at admission, at the peak of viral load (Figure 1a and 1b), or during the course of the disease (Figure 2). Recipient age (ρ=0.23, p=0.16, Figure 3) and sex (p=0.05) were marginally associated with the viral load. Notably, patients receiving steroid therapy and not presenting gastrointestinal symptoms displayed a higher viral load (Table 2). No correlation was evident between maximum viral load and inflammatory markers, including interleukin (IL)-6 (ρ=0.041, p=0.81) and C-reactive protein (CRP, ρ=0.001, p=0.99). Among patients with an initial positive RT-PCR test result (n=37), no patient showed a viral clearance before D21. Fifteen (38.5%) patients displayed a positive viral load greater than 3 log^10^ copies/reaction after D10, and ten patients (24.4 %) showed persistent viral shedding after D30 (Figure 2).

**Table 2:** SARS-CoV 2 nasopharyngeal viral load in log_10_ copies/reaction according to demographic and clinical characteristics of hospitalized kidney transplant recipients (N=39)

|  | **n** | **Nasopharyngeal maximal viral load** | **p** |
| --- | --- | --- | --- |
| **COVID-19 severity** |  |  | **0.23** |
| **Non-severe disease** | 21 | 5.17 [3.80-6.61] |  |
| **Severe disease** | 18 | 6.38 [4.88-7.21] |  |
| **Recipient age, years** |  |  | **0.70** |
| **< 60** | 24 | 5.26 [3.42-6.92] |  |
| **≥ 60** | 15 | 5.88 [4.43-7.04] |  |
| **Sex** |  |  | **0.05** |
| **Female** | 8 | 7.34 [5.74-8.02] |  |
| **Male** | 31 | 5.17 [4.32-6.63] |  |
| **BMI, kg/m<sup>2</sup>** |  |  | **0.08** |
| **<30** | 20 | 6.50 [5.10-7.32] |  |
| **≥30** | 19 | 4.90 [3.43-6.42] |  |
| **Obstructive sleep apnea** |  |  | **0.57** |
| **No** | 32 | 5.81 [4.71-7.14] |  |
| **Yes** | 7 | 4.44 [3.50-4.44] |  |
| **Cardiovascular disease** |  |  | **0.84** |
| **No** | 23 | 6.01 [4.88-6.84] |  |
| **Yes** | 16 | 5.04 [3.80-7.50] |  |
| **Respiratory disease** |  |  | **0.31** |
| **No** | 24 | 6.26 [4.62-7.28] |  |
| **Yes** | 15 | 4.90 [4.32-6.70] |  |
| **Diabetes** |  |  | **0.11** |
| No | 21 | 6.01 [4.90-7.55] |  |
| Yes | 18 | 5.04 [3.24-6.45] |  |
| **Hypertension** |  |  | **0.84** |
| No | 7 | 5.17 [4.35-6.82] |  |
| Yes | 32 | 5.81 [4.37-7.14] |  |
| **RAAS inhibitor** |  |  | **0.17** |
| No | 25 | 6.38 [4.90-7.11] |  |
| Yes | 14 | 6.88 [3.34-6.53] |  |
| **Immunosuppressive induction therapy** |  |  | **0.20** |
| Anti-thymocyte globulin | 18 | 6.38 [2.59-7.61] |  |
| Anti-CD25 | 18 | 5.17 [3.96-6.45] |  |
| No induction | 3 | 4.90 [3.85-6.07] |  |
| **Immunosuppressive maintenance therapy** |  |  |  |
| **CNI** |  |  | **0.71** |
| No | 5 | 4.90 [4.42-7.76] |  |
| Yes | 34 | 5.81 [3.96-7.00] |  |
| **MMF/MPA** |  |  | **0.15** |
| No | 6 | 7.08 [6.34-7.81] |  |
| Yes | 33 | 5.17 [4.21-6.66] |  |
| **mTORi** |  |  | **0.92** |
| No | 33 | 5.61 [4.21-7.02] |  |
| Yes | 6 | 5.92 [4.54-7.16] |  |
| **Steroids** |  |  | **0.03** |
| No | 17 | 4.87 [3.05-6.38] |  |
| **Yes** | 22 | 6.50 [5.17-7.53] |  |
| **Clinical symptoms** |  |  |  |
| **Dyspnea** |  |  | **0.23** |
| **No** | 12 | 4.79 [2.73-6.71] |  |
| **Yes** | 27 | 6.15 [4.88-7.17] |  |
| **Diarrhea** |  |  | **0.02** |
| **No** | 9 | 6.93 [6.38-7.41] |  |
| **Yes** | 30 | 5.17 [3.80-7.83] |  |
| **Positive RNAemia** |  |  | **0.35** |
| **No** | 18 | 5.17 [4.26-6.88] |  |
| **Yes** | 9 | 6.61 [5.26-7.24] |  |

Nasopharyngeal maximal viral load are presented as median (interquartile range)
Abbreviations: BMI, body mass index; RAAS, renin-angiotensin-aldosterone system; MMF, mycophenolate mofetil; MPA, mycophenolic acid, mTOR: mammalian target of rapamycin.

**Figure 1:**
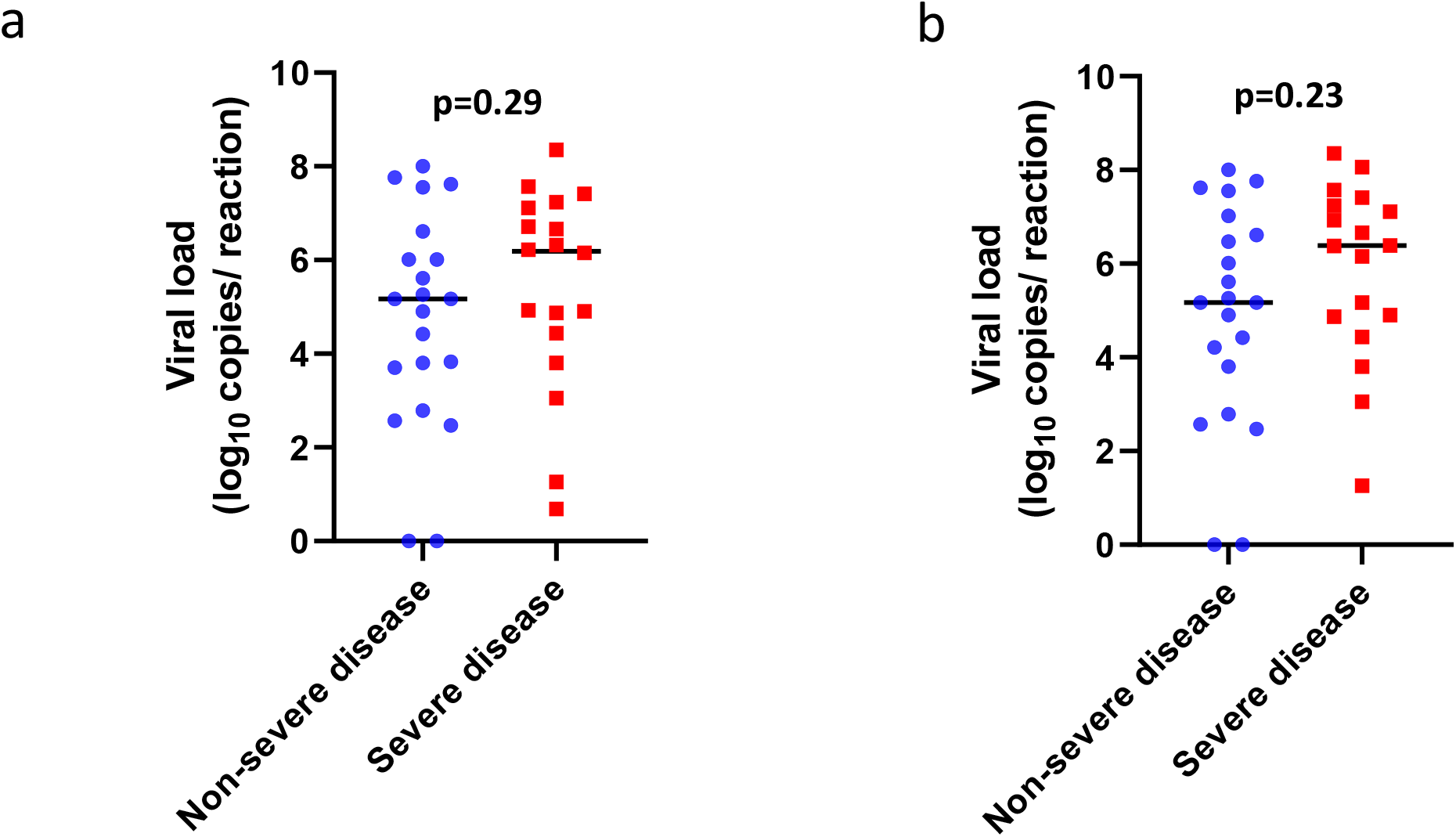
SARS-CoV-2 viral load distribution according to disease severity in nasopharyngeal swab. A: scatter plots with the medians (black lines) of the viral loads at admission in non-severe (blue) and severe patients (red). B: scatter plots with the medians (black lines) of the maximum viral loads during the follow-up in non-severe (blue) and severe patients (red).

**Figure 2:**
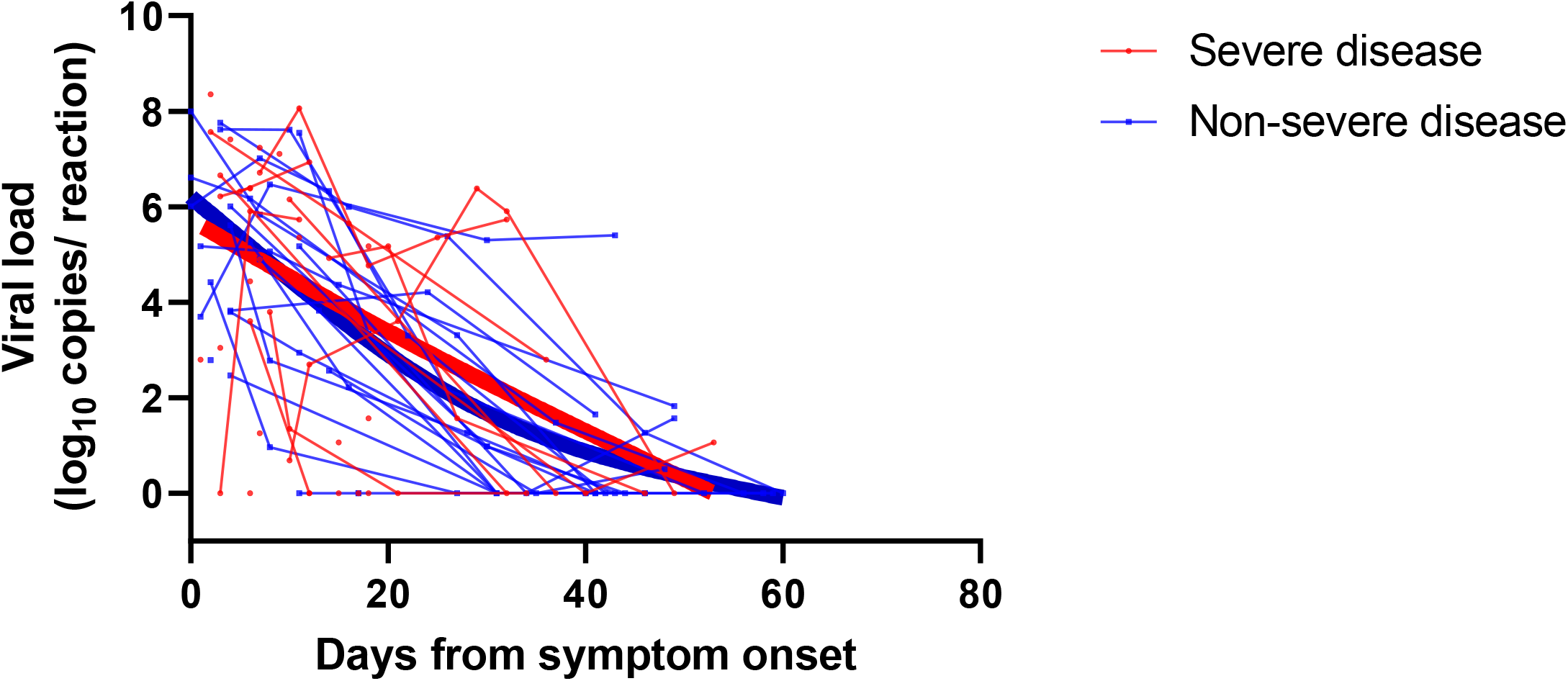
SARS-CoV-2 viral load kinetics analyzed using nasopharyngeal swabs. Patients are stratified according to non-severe disease (blue) and severe disease (red). The thick lines show the trend in viral load using smoothing splines.

**Figure 3:**
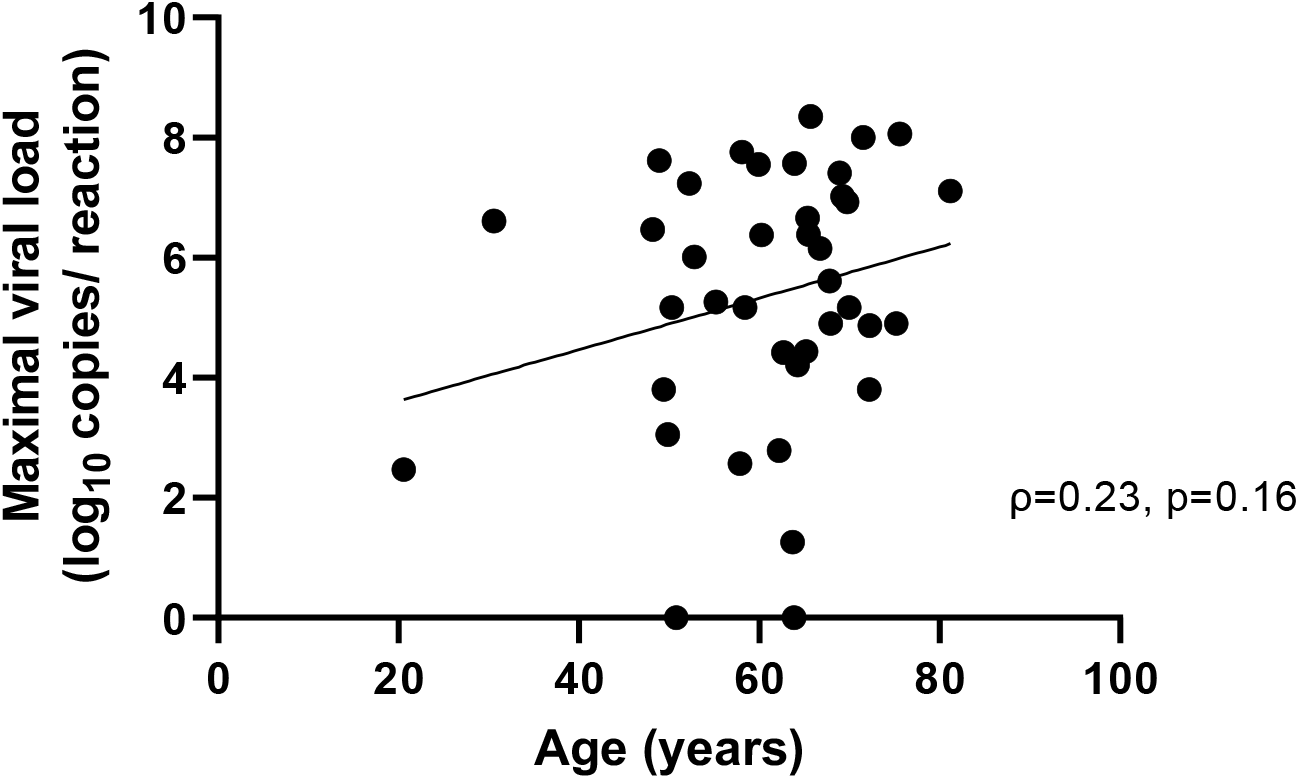
Spearman’s correlation between the age of a kidney transplant recipient and the maximum viral load determined using a nasopharyngeal swab (Spearman’s correlation coefficient ρ=0.23, p=0.16).

### 3.3 SARS-CoV-2 RNAaemia

SARS-CoV-2 loads were measured in 73 plasma samples obtained from 32 patients (21 in the non-severe group and 11 in the severe group). Plasma viral loads ranged from 1 to 4.55 log^10^ copies/reaction. Ten patients had at least one positive RNAaemia. Severe patients showed a higher frequency of RNAaemia compared with non-severe patients (50% *versus* 26.3%, respectively, p=0.0087, Figure 4a). Moreover, RNAaemia was found to be associated with mortality (Figure 4b). Accordingly, RNAaemia was positive in all three non-survivors tested; in contrast, only 7 of 29 (24%) tested survivors had positive RNAaemia (p=0.024).

**Figure 4:**
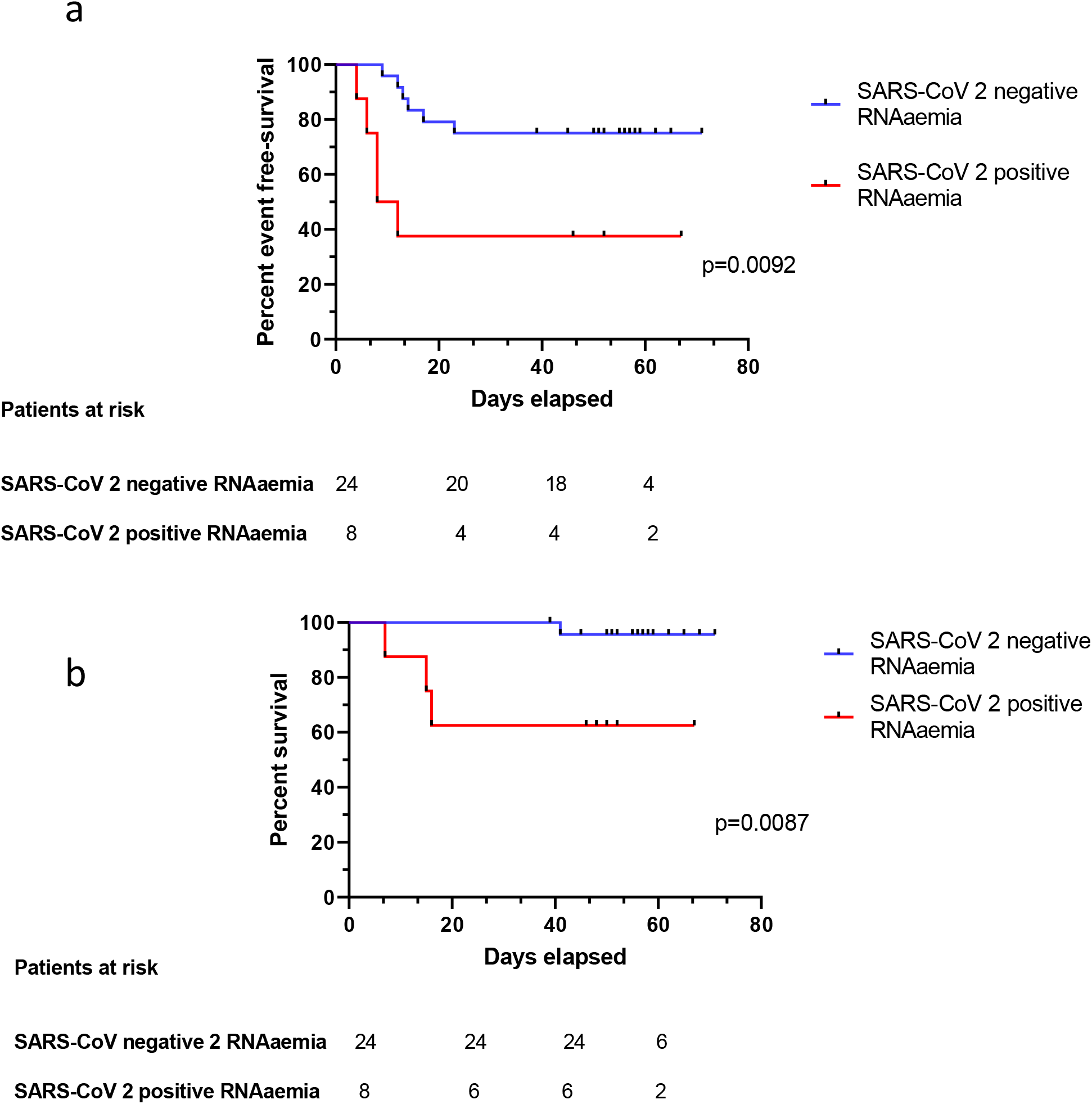
Association of positive SARS-CoV-2 RNAaemia with COVID-19 severity and mortality. A. Kaplan-Meier plots of COVID-19-free survival according to SARS-CoV-2 RNAemia. Presence of SARS-CoV-2 RNAaemia (red curve) *versus* its absence (blue curve), p=0.0092. B. Kaplan-Meier plots of severe COVID-19-free survival according to SARS-CoV-2 RNAemia. Presence of SARS-CoV-2 RNAaemia (red curve) *versus* its absence (blue curve), p=0.0087.

Furthermore, two non-survivors harbored high viral loads (4.55 log^10^ copies/reaction and 4.26 log^10^ copies /reaction), whereas other patients were characterized by low RNAaemia (< 2.16 log^10^ copies/reaction). With regard to immunosuppressive therapy, patients receiving CNI tended to have more positive RNAaemia (10/24 *versus* 0/5, respectively, p=0.13).

### 3.4 SARS-CoV-2 serological findings

A total of 116 samples from 35 patients were analyzed, with a median of three sera tested per patient (IQR: 2−4 sera). All survivors were seropositive at follow-up. Four non-survivors had negative serology at their time of death, which occurred on D7, D10, D15, and D16. The two patients with a negative SARS-CoV-2 RT-PCR result had positive serology, with one patient showing positive serology at the time of diagnosis. Among the 25 samples tested before D8, six (24%) were seropositive, whereas all samples tested after D14 were seropositive (Figure 5). The kinetics of the antibodies showed that a stable titer of IgG antibodies was maintained until D59, suggesting persistence of immunity for at least until two months after infection (Figure 6). Notably, IgM and IgG antibody level and delays in seroconversion were not correlated with COVID-19 severity (Figure 7a and 7b).

**Figure 5:**
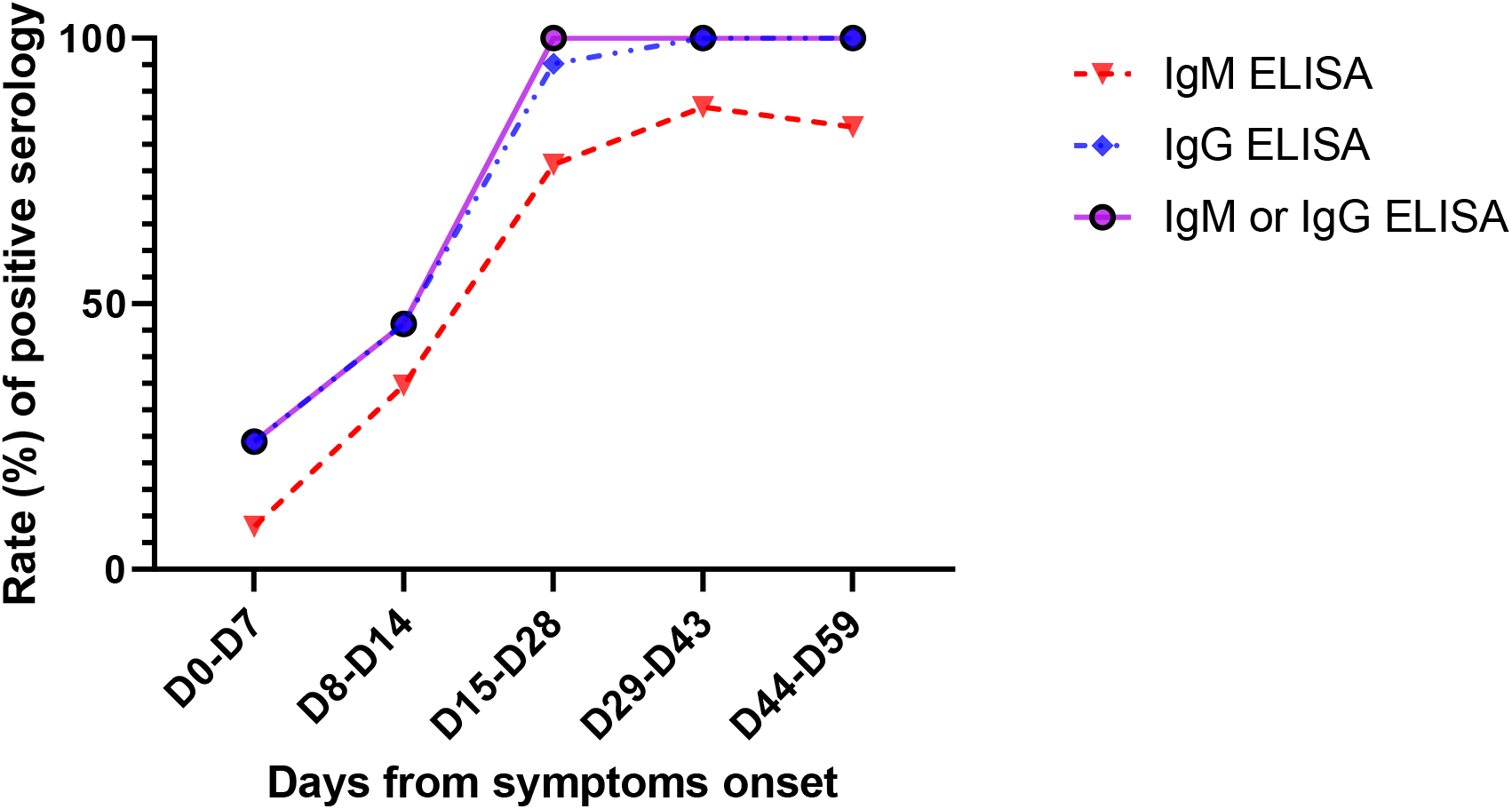
The rate of positive SARS-CoV-2 IgM (red curve), IgG (blue curve), and IgM or IgG (purple curve) tested by an enzyme-linked immunosorbent assay (ELISA) according to the days from symptom onset. A total of 116 samples from 35 patients were tested. From day 15 (D15) onwards, all samples were positive for IgM or IgG.

**Figure 6:**
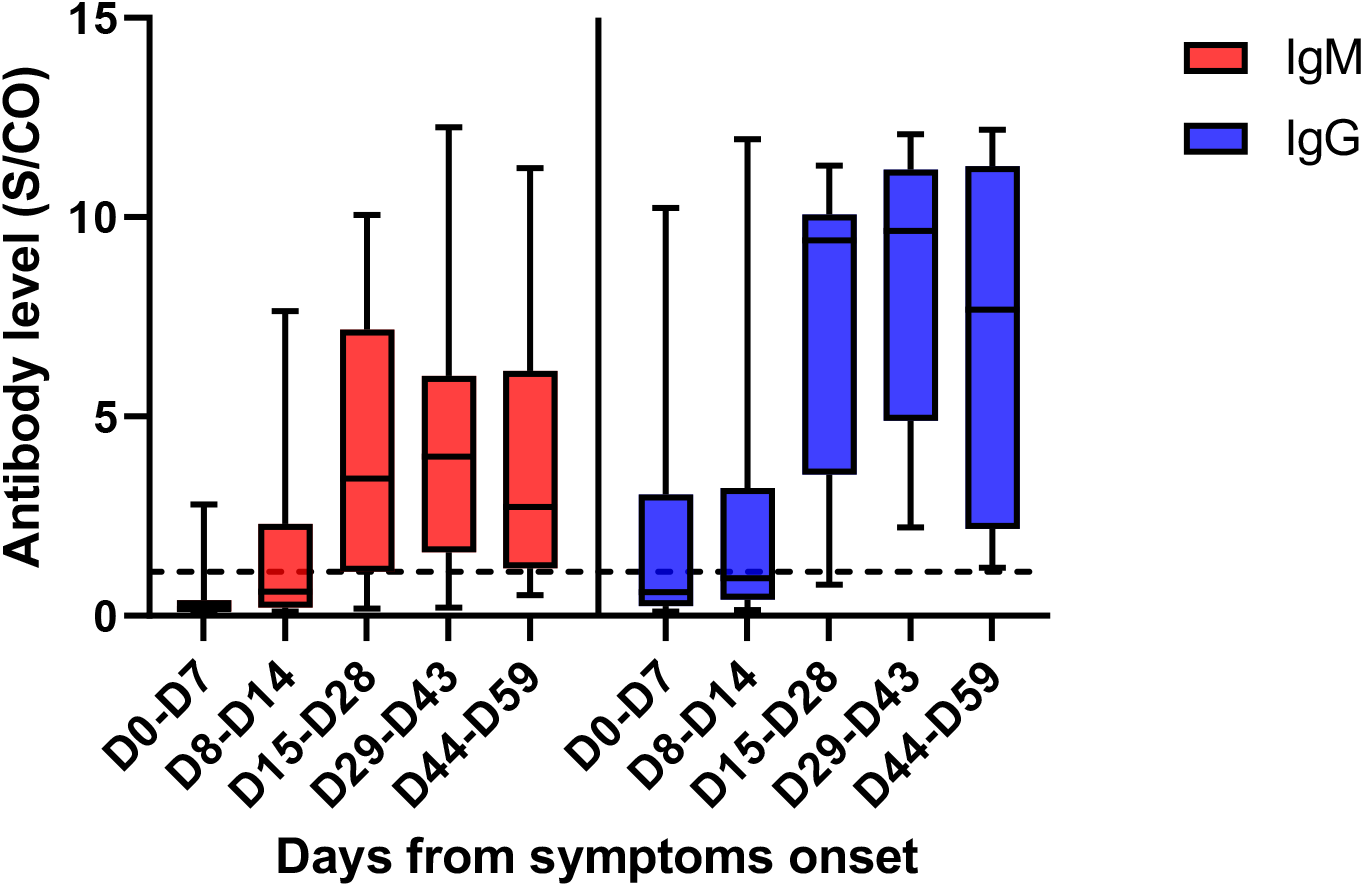
SARS-CoV-2 IgM (red plots) and IgG (blue plots) titers tested by an enzyme-linked immunosorbent assay (ELISA), according to the days from symptom onset. IgM and IgG levels increased significantly over time. Antibody levels are presented as the measured absorbance values divided by the cutoff (S/CO). The dotted line represents the cutoff value (1.1). The boxplots show medians (middle line) and first and third quartiles (boxes), whereas the whiskers indicate minimum and maximum values.

**Figure 7:**
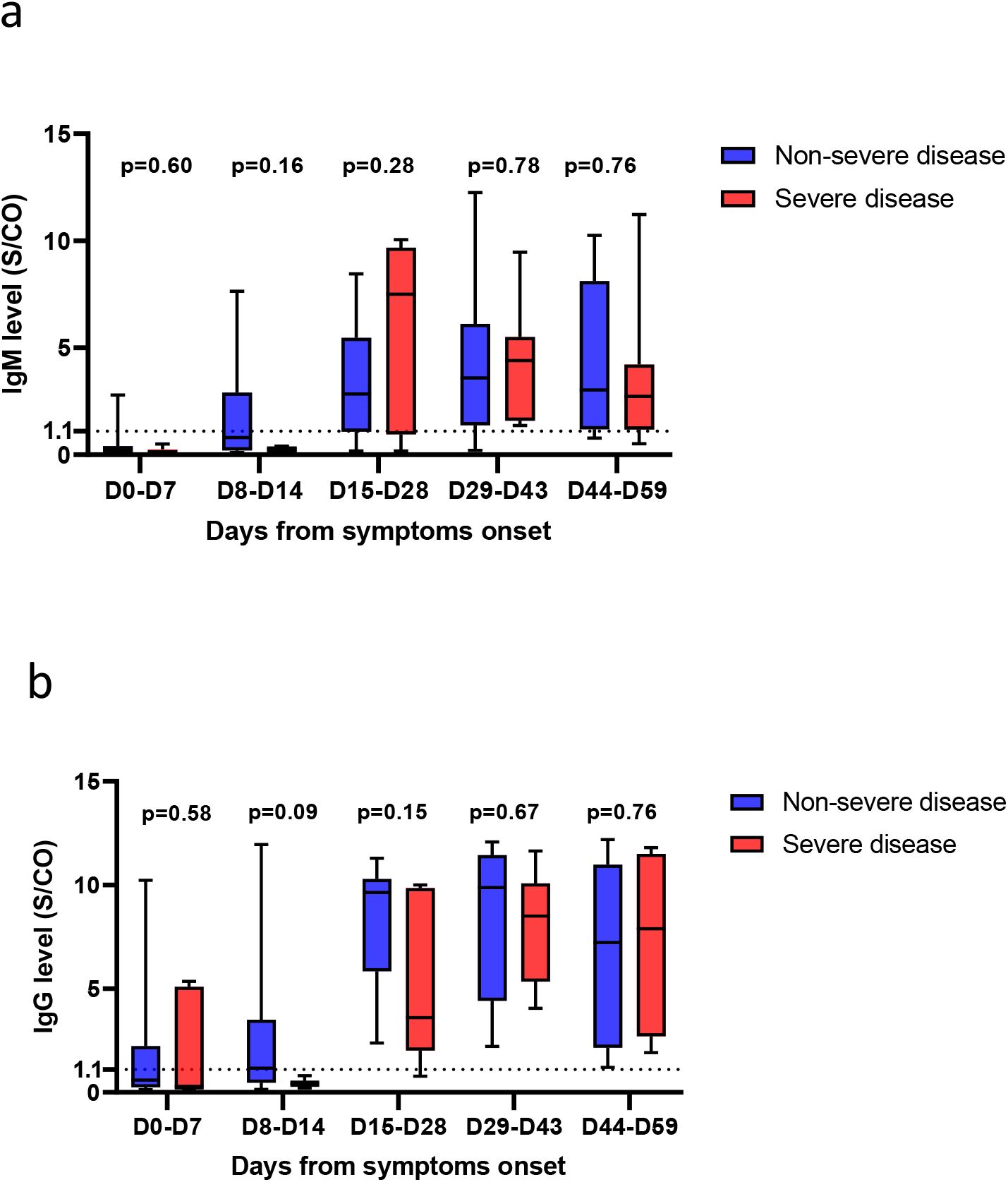
SARS-CoV-2 IgM (Figure 6a) and IgG (Figure 6 b) titers tested by an enzyme-linked immunosorbent assay (ELISA) according to the days from symptom onset and stratified by disease severity (severe [red] *versus* non-severe [blue]). IgM and IgG antibody levels did not differ according to disease severity (p>0.05). Antibody levels are presented as the measured absorbance values divided by the cutoff (S/CO). The dotted line represents the cutoff value (1.1). The boxplots show medians (middle line) and first and third quartiles (boxes), whereas the whiskers indicate minimum and maximum values.

## 4. Discussion

In this retrospective study conducted in a sample of 40 immunocompromised KTR hospitalized for COVID-19, we precisely determined the temporal evolution of nasopharyngeal and plasma SARS-CoV-2 loads, as well as the serological response to the virus. All parameters were correlated with patient characteristics, disease severity, and clinical outcomes. In our study, the viral load in the respiratory specimens of most patients was at the peak at the time of diagnosis. This finding is in line with those previously reported for the general population.^5–7^ Based on the data we analyzed, the viral load at the time of diagnosis did not predict the severity of the disease. Moreover, viral loads were not related to inflammatory markers that have been previously associated with COVID-19 severity.^8^ Reports on the relationship between viral load and disease severity are contradictory; three studies showed no correlation between the severity of the disease and the viral load in respiratory specimens,^6,7,9^ whereas Liu et al.^10^ described a higher viral load in patients with more severe disease.

In our immunocompromised population with a median follow-up of 53 days, the duration of viral detection in the respiratory tract was longer compared to that in the general population. More than a third of our patients displayed a high viral load after D10. Furthermore, almost a quarter of them still had viral shedding at D30; in contrast, in immunocompetent populations, the median duration of viral shedding was 20 days.^12^ Thus, even if a positive RT-PCR result does not indicate an infectious virus, we should be cautious about viral spread in vulnerable populations and extend isolation after a SARS-CoV-2 infection.

SARS-CoV-2 RNAaemia was positive in 32% of our KTR with COVID-19. To our knowledge, SARS-CoV-2 RNAaemia in immunocompromised patients had not been previously described. Nonetheless, this finding is in accordance with what was observed in the general population (ranging from 10.4% to 41% positivity for RNAaemia).^8,12,13^ Our study shows that RNAaemia is associated with disease severity and mortality in KTR patients.

However, published data on the subject are mixed. In studies conducted by Huang et al.^12^ and Zheng et al.^13^, RNAaemia was not associated with COVID-19 severity, whereas in the cohort study carried out by Chen et al.^8^ positive RNAaemia was correlated with highly elevated IL-6 plasma leveland was only detected in critically ill patients. In the study by Hadjadj et al.^9^, patients with severe and critical COVID-19 had higher plasma viral loads than those with mild and moderate disease. The pathophysiological mechanism of the association between COVID-19 severity and plasma viral load is still unclear; perhaps the cytokine storm that affects COVID-19 severity could enhance RNAaemia by causing increased vascular permeability, or a significant viral load could trigger the cytokine storm.^8^

Regarding the SARS-CoV-2 humoral response, this is the largest study with the longest follow-up in an immunocompromised population. All but four patients had positive serology during the follow-up. The four negative patients were tested early in the course of the disease and died shortly after. A total of 13 (43.3%) patients displayed positive serology before D15, supporting the potential usefulness of serology tests for acute diagnosis. All patients harbored SARS-CoV-2 antibodies from D15 onwards. Data on the serological response to SARS-CoV-2 in immunocompromised populations remain scanty. In a study from the United States, nine patients infected with SARS-CoV-2 after a solid organ transplant had positive IgG serology, with a delay of seroconversion between D6 and D27.^14^ Zhao et al.^15^ reported a delayed antibody response in a patient co-infected with COVID-19, HIV, and hepatitis C. In the general population, studies have reported seroconversion for all patients between the third and fourth week after symptom onset, which is concordant with our findings. These results suggest that the anti-SARS-CoV-2 humoral response is not significantly impaired in our immunocompromised population.^16, 17, 18^ Notably, the delay after transplantation was long in our cohort, and only one patient had undergone depleting induction therapy during the year preceding COVID-19. The level of IgG remained steady until two months after onset of symptoms. Although the neutralizing effect of the antibodies was not studied here, it is encouraging that IgG levels are correlated with a neutralizing effect in the general population.^6,16^

Despite some limitations of this study, including the small sample size and lack of data from some patients, this is the first report that provides a precise assessment of SARS-CoV-2 virological and antibody response kinetics in an immunocompromised population, with a follow-up for two months after symptom onset. Taken together, our data indicate that 1) SARS-CoV-2 shedding from the upper respiratory tract is prolonged in KTR patients, indicating the requirement for prolonged protective measures for these patients; 2) the SARS-CoV-2 plasma load is associated with COVID-19 severity and mortality, whereas the viral load of the upper respiratory tract is not; and 3) based on the presence of antibodies in all samples collected by us after the second week of symptom onset, the SARS-CoV-2 humoral response in our immunocompromised population does not show serious impairment and the antibodies persist until two months after COVID-19 symptom onset.

## Data Availability

Data supporting the findings of this study are available from the corresponding author upon reasonable request.

## Abbreviations

AUC: area under curve
BMI: body mass index
CNI: calcineurin inhibitor
COVID-19: coronavirus disease-2019
CRP: C-reactive protein
CT: computed tomography
D: day after symptom onset
ELISA: enzyme-linked immunosorbent assay
ICU: intensive care unit
Ig: immunoglobulin
IL: interleukin
KTR: kidney transplant recipients
MMF: mycophenolate mofetil
MPA: mycophenolic acid
mTOR: mammalian target of rapamycin
RdRp: *RNA-dependent RNA polymerase*
ROC: receiver operating characteristic
RT-PCR: reverse transcription-polymerase chain reaction
SARS-CoV-2: severe acute respiratory syndrome coronavirus-2
S/CO: absorbance values divided by the cutoff
WHO: World Health Organization

## Acknowledgments/funding

this study was supported by the Strasbourg University Hospital (COVID-HUS study-HUS N°7760).

